# Impact of blood analysis and immune function on the prognosis of patients with COVID-19

**DOI:** 10.1101/2020.04.16.20067587

**Authors:** Yue-qiang Fu, Yue-lin Sun, Si-wei Lu, Yang Yang, Yi Wang, Feng Xu

## Abstract

**Introduction:** This retrospective study investigated the implications of changes in blood parameters and cellular immune function in patients with 2019-coronavirus infected disease (COVID-19).

**Methods:** Records were reviewed of 85 patients with COVID-19 between February 4 and 16, 2020. The primary outcome was in-hospital mortality at 28 days.

**Results:** Fourteen patients died. The baseline leukocyte count, neutrophil count and hemoglobin was significantly higher in non-survivors compared with survivors, while the reverse was true of lymphocyte count, platelet, PaO_2_/FiO_2_, CD3+ count and CD4+ count. The percentage of neutrophil count > 6.3×10^9^/L in death group was significantly higher than that in survival group, and multivariate logistic regression showed neutrophil count was independently associated with mortality. However, there were not significant difference in IgG, IgM, IgA, C3, C4 and the percentage of IgE > 100 IU/ml between the death group and survival group. Areas under the receiver operating characteristic curves of the following at baseline could significantly predict mortality: leukocyte, neutrophil, lymphocyte, CD3+ and CD4+ counts.

**Conclusions:** For patients with COVID-19, lymphocyte, CD3+ and CD4+ counts that marked decrease suggest a poor outcome. A high neutrophil count is independently associated with mortality. At admission, leukocyte, neutrophil, lymphocyte, CD3+ and CD4+ counts should receive added attention.

## INTRODUCTION

The first case of an unknown pneumonia was traced to the Wuhan South China Seafood Market in December 2019. It was confirmed as an acute respiratory infectious disease caused by severe acute respiratory syndrome coronavirus 2 (SARS-CoV-2, formerly known as 2019-nCoV) and disease has been subsequently named 2019-coronavirus infected disease (COVID-19) by WHO on Feb 11, 2020 [1]. At present, the COVID-19 epidemic has wrought great damage in China, Asia, Europe, and North America, and globally all countries are under threat. COVID-19 is highly contagious and can cause serious lung injury, resulting in death. Medical research concerning this new virus is relatively limited, and the pathophysiological mechanisms underlying the cause of disease are still under study.

Pulmonary infection caused by virus is often accompanied by changes in hematologic and immune test parameters. Neutrophil, lymphocyte, and lymphocyte subset counts may help clinicians assess the condition of patients with viral pneumonia. In patients with severe acute respiratory syndrome (SARS), an initial neutrophil count >7000/mL (odds ratio [OR] 6.4) and initial C-reactive protein (CRP) concentration >47.5 mg/L (OR 5.8) have been associated with fatality [2]. Leong et al. [3] found that neutropenia was associated with poor prognosis in patients with SARS, and Li et al. [4] reported that the peripheral CD4+ and CD8+ T lymphocyte absolute counts were significantly lower in these patients. Tang et al. [5] also showed that CD3+, CD4(+), and CD8(+) lymphocytes significantly decreased in the acute phase of SARS, especially in patients who died.

Wang et al. [6] showed that most patients with COVID-19 had marked lymphopenia, and non-survivors developed more severe lymphopenia over time during hospitalization. Another study also indicated that COVID-19 patients in the death group had significantly lower lymphocyte count on admission than the recovered group [7], however they did not explore changes in lymphocyte subsets. Recently, several studies showed that compared to non-severe cases, the severe COVID-19 cases had lower lymphocyte counts [8-13] and higher leukocytes counts [8-10]. Wang et al. [13] found that CD4+ T cells, CD8+ T cells, B cells and natural killer (NK) cells decreased in COVID-19 patients, and severe cases had a lower level than mild cases. Other studies also mentioned that CD4+T and CD8+T cells decreased in the COVID-19 patients [8,14]. However, whether there are changes in humoral immune function has not been studied yet. And the differences of lymphocyte subsets between the survival and the death patients with COVID-19 are rarely studied.

COVID-19 may lead to hyperactivity of the inflammatory response, and suppression of the immune response. Changes in leukocyte subsets and immune function after SARS-CoV-2 infection may suggest immune status, but this is not well studied. The objective of the present case series was to compare the clinical characteristics, blood analyses, and immune functions features between the survivors and non-survivors. The predictive value of neutrophil, lymphocyte, and lymphocyte subsets at admission for mortality was also studied.

## PATIENTS AND METHODS

### Study design

The Ethics Committee of Children’s Hospital, Chongqing Medical University (Institutional Review Board of Children’s Hospital, Chongqing Medical University) approved this retrospective cohort study. The requirement for written informed consent was waived because of the retrospective design.

The study population comprised patients with COVID-19 pneumonia, who had been admitted to the ward of the Third Batch of Chongqing Medical Aid Team in Wuhan city of Hubei province in China, from 4 February 2020 to 16 February 2020. The data were analyzed anonymously.

The patients in this study were confirmed to have SARS-CoV-2 infection by reverse real-time reverse transcription polymerase chain reaction (RT-PCR) assay, from nose and throat swab samples. At least one ground glass change in the lung was indicated on chest computed tomography (CT) scan. A blood routine and immune function test was completed within 12 hours after admission. Patients with any of the following were excluded from this study: death due to natural causes rather than viral infection; immunodeficiency; or with long-term use of glucocorticoids or immunosuppressants.

### Data collection and definitions

Collected information included: age; gender; concomitant disease; symptoms; hemoglobin; platelet; leukocyte, neutrophil, and lymphocyte counts; CRP, CD3+, CD4+, CD8+, CD4/8, CD19+, and CD16+56+;IgG, IgM, IgA, IgE, C3, and C4 chest CT; Alanine aminotransferase, Creatinine; arterial blood gas; and the result of the RT-PCR assay of SARS-CoV-2 RNA.

### Outcome

Patient outcomes (survival or death) were followed until 16 March 2020. The primary outcome was in-hospital mortality at 28 days.

### Statistical analysis

Data were analyzed using SPSS 21.0 (SPSS, Chicago, IL, USA). Continuous variables are shown as median (interquartile range or IQR), and categorical data as percentage. The comparison of two medians was performed using the Mann-Whitney U test; and the comparison of percentages with the chi-squared test. Multivariate analysis was performed to determine whether the admission neutrophil count was a risk factor for mortality, after adjusting for admission lymphocyte count, PaO_2_/FiO_2_, CD3+, CD4+ and CRP >60 mg/L. Receiver operating characteristic (ROC) curves were used to determine blood parameters at admission as predictors of mortality. A *P*-value < 0.05 was considered statistically significant for all analyses.

## RESULTS

### Clinical characteristics

Overall, 93 persons were treated during the study period, 85 (49 men) of whom met the inclusion criteria for this study (Table 1). The remaining 8 others were excluded due to absence of specific blood test within 12 hours after admission, negative SARS-CoV-2 result of nasopharyngeal swab sample, or death due to advanced age rather than virus infection. The age of the study population was 64.00 (54.50, 70.00) years (range 31-89 y). The interval between symptom onset and admission was 13.00 (10.00,15.00) days. Of these 85 patients, 44 suffered from concomitant chronic diseases. There was at least one ground glass change in the chest CT scans of each of these patients. As of 16 March 2020, 46 patients were discharged, 14 had died, and the remaining 25 were still hospitalized. Median hospitalization days of patients who died was 4.00 (2.75, 6.25) days. The most common clinical symptoms are fever and cough. For this analysis, the 85 patients were assigned either to the survival group or the death group.

**Table 1.** Demographics, clinical characteristics, and blood indexes of the survival and death groups

|  | All patients | Survival group | Death group | *P* |
| --- | --- | --- | --- | --- |
| Subjects, n (%) | 85 | 71 | 14 | — |
| Age, y | 64.00(54.50, 70.00) | 62.00(55.00, 70.00) | 67.00(50.75, 74.25) | 0.495 |
| Gender, male, n (%) | 49 (57.65%) | 38(53.52%) | 11(78.57%) | 0.083 |
| Any comorbidity, n (%) | 47(55.29%) | 37(52.11%) | 10(71.43%) | 0.184 |
| Hypertension, n (%) | 31(36.47%) | 24(33.80%) | 7(50.00%) | 0.250 |
| Coronary heart disease, n (%) | 12(14.12%) | 8(11.23%) | 4(28.57%) | 0.089 |
| Diabetes, n (%) | 13(15.29%) | 9(12.68%) | 4(28.57%) | 0.131 |
| Between onset and hospitalization, d | 13.00(10.00,15.00) | 13.00(10.00, 15.00) | 12.00(9.00,15.25) | 0.487 |
| Fever, n (%) | 77(90.59%) | 64(90.14%) | 13(92.86%) | 0.750 |
| Cough, n (%) | 66(77.65%) | 55(77.46%) | 11(78.57%) | 0.928 |
| Dyspnea, n (%) | 34(40%) | 22(30.99%) | 12(85.71%) | 0.000 |
| Diarrhea, n (%) | 19(22.35%) | 17(23.94%) | 2(14.29%) | 0.455 |
| Chest tightness, n (%) | 45(52.94%) | 35(49.30%) | 10(71.43%) | 0.107 |
| Fatigue, n (%) | 41(48.24%) | 34(47.89%) | 7(50%) | 0.885 |
| PaO<sub>2</sub>/FiO<sub>2</sub> | 213.51(170.27, 259.46) | 218.92(191.89, 270.27) | 128.38(98.65, 191.89) | 0.000 |
| Leukocyte count ×10<sup>9</sup>/L | 6.26(4.52, 8.64) | 5.71(4.33, 7.39) | 11.20 (7.59, 14.60) | 0.000 |
| Neutrophil percentage % | 73.20(65.10, 84.65) | 69.30(62.10, 79.10) | 89.65 (85.63, 92.28) | 0.000 |
| Neutrophil count ×10<sup>9</sup>/L | 4.55(3.00, 6.67) | 3.96(2.85, 5.72) | 10.10(6.58, 13.49) | 0.000 |
| >6.3, n (%) | 24(28.24%) | 12(16.90%) | 12(85.71%) | 0.000 |
| Lymphocyte percentage % | 17.70(8.45, 23.35) | 20.30(11.80, 24.80) | 5.30(4.05, 7.80) | 0.000 |
| Lymphocyte count ×10<sup>9</sup>/L | 0.96(0.62, 1.42) | 1.00 (0.77, 1.43) | 0.57 (0.42, 0.80) | 0.002 |
| < 1.1, n (%) | 53(62.35%) | 41(57.75%) | 12(85.71%) | 0.048 |
| Hemoglobin g/L | 111.50(122.00, 135.50) | 121.00(111.00, 132.00) | 140.00(121.00, 156.25) | 0.017 |
| Platelets ×10<sup>9</sup>/L | 214.00(161.00, 270.00) | 224.00(179.00, 275.00) | 161.00(136.75, 198.25) | 0.004 |
| CRP > 60 mg/L, n (%) | 32(37.65%) | 21(29.58%) | 11(78.57%) | 0.001 |
| Alanine aminotransferase, U/L | 29.00 (20.00, 55.00) | 29.00 (19.00, 51.00) | 41.00 (22.50, 88.50) | 0.361 |
| Creatinine, μmol/L | 55.00 (47.00, 69.50) | 55.00 (46.00, 69.00) | 67.50 (48.75, 74.00) | 0.361 |

Data shown are number (%) or median (interquartile range). CRP- C-reactive protein; FiO<sub>2</sub>- Fraction of inspiration O<sub>2</sub>; PaO<sub>2</sub>- partial pressure of oxygen.

There was no significant difference in age between the survival and death groups [62.00(55.00, 70.00) y cf. 67.00(50.75, 74.25) y; Table 1]. In the death group, the percentage of men (11/14, 78.6%) and rate of accompanying disease (10/14, 71.4%) were higher compared with the survival group (38/71, 53.52%; and 37/71, 52.11%, respectively), but the differences were not significant. The incidence of dyspnea in the death group was higher than that in the survival group. The survival and death groups were also comparable regarding the interval between onset and admission [13.00 (10.00,15.50) d cf. 12.00 (9.00,15.25) d].

### Blood analysis parameters in survivors and non-survivors

The following blood parameters in the death group were significantly higher than in the survival group (Table 1): total leukocyte count (*P* = 0.000); percentage of neutrophils (*P* = 0.000), neutrophil count (*P* = 0.000) and hemoglobin (*P* = 0.017). Upon admission, the percentage of the death group with CRP >60 mg/L was significantly higher than that of the survival group [(21/71 (29.58%) cf. 11/14 (78.6%)]. In addition, 12/14 (85.71 %) patients in the death group had an admission neutrophil count > 6.3 × 10^9^/L, while this was true of only 12/71 (16.90%) of the survival group. In the death group, the percentage of lymphocytes, lymphocyte count, platelet count, and PaO_2_/FiO_2_ were significantly lower than that of the survival group (*P* = 0.000, 0.002, 0.004, 0.000, respectively). 12/14 (85.7%) patients in the death group had lymphocyte count at baseline < 1.1 × 10^9^/L, however this was true of 41/71 (57.75%) of the survival group.

### Cellular and humoral immunity in survivors and non-survivors

In the 85 patients, the baseline median of the CD3+, CD4+, and CD8+ counts were all lower than the lower limit of the normal range (Table 2). The CD3+, CD4+ and CD8+ counts in more than 60% of patients were lower than the lower limit of normal range, respectively; the incidence rates were higher in non-survivors than survivors (Table 2). The median CD3+%, CD3+ count, CD4+%, and CD4+ count in the death group were all significantly lower than that of the survival group (*P* = 0.020, 0.007, 0.006 and 0.001, respectively). The medians of the CD8+ count and CD16+56+ count, CD19+ count and CD4+/CD8+ ratio in the death group were lower than those of the survival group, but the differences were not statistically significant.

**Table 2.** Cellular immunity in survival and death groups

|  | Normal range | All patients | Survival | Death | *P* |
| --- | --- | --- | --- | --- | --- |
| Subjects, n |  | 85 | 71 | 14 | — |
| CD3+ percentage, % | 56-86 | 65.16 (53.92, 74.03) | 69.00 (55.48, 74.82) | 60.49 (45.30, 64.45) | 0.020 |
| CD3+ count, /μL | 723-2737 | 558.00 (345.50,818.50) | 609.00 (410.00,905.00) | 339.50 (217.50, 524.25) | 0.007 |
| <723 /μL, n (%) | | 58 (68.24%) | 45 (63.38%) | 13 (92.86%) | 0.030 |
| CD4+ percentage, % | 33-58 | 40.64 (32.39, 45.56) | 41.46 (32.95, 46.58) | 33.90 (27.09, 39.58) | 0.006 |
| CD4+ count, /μL | 404-1612 | 326.00 (202.50, 479.00) | 368.00 (246.00, 549.00) | 203.00 (126.50, 284.25) | 0.001 |
| <404 /μL, n (%) | | 54 (63.53%) | 40 (56.34%) | 14 (100%) | 0.002 |
| CD8+ percentage, % | 13-39 | 21.95 (15.92, 30.10) | 22.20 (16.52,29.90) | 20.65 (13.36,32.06) | 0.648 |
| CD8+ count, /μL | 220-1129 | 191.00 (99.00, 298.00) | 205.00(111.00, 303.00) | 145.00 (70.00, 213.00) | 0.067 |
| <220, /μL, n (%) | | 52 (61.18%) | 40(56.34%) | 12(85.71%) | 0.039 |
| CD4+/CD8+ ratio | 0.9-2.0 | 1.82 (1.23,2.67) | 1.93 (1.26,2.68) | 1.59 (1.13, 2.47) | 0.554 |
| <0.9, n (%) |  | 7 (8.24%) | 5 (7.04%) | 2 (14.29%) | 0.368 |
| CD19+ percentage, % | 5-22 | 15.50(11.29,22.20) | 15.23(11.55,21.22) | 17.30(10.33, 40.30) | 0.238 |
| CD19+ count, /μL | 80-616 | 123.00(88.50,171.50) | 128.00(91.00,187.00) | 106.00(55.00,142.75) | 0.081 |
| <80, /μL, n (%) | | 14(16.47%) | 10(14.08%) | 4(28.57%) | 0.182 |
| CD16+56, % | 5-26 | 13.37(9.20,21.43) | 13.24(8.79,19.03) | 17.32(11.59,26.48) | 0.159 |
| CD16+56 count, /μL | 84-724 | 118.00(68.50, 170.50) | 119.00(74.00, 171.00) | 88.00(39.5, 176.25) | 0.470 |
| <84, /μL, n (%) | | 29(34.12%) | 23(32.39%) | 6(42.86%) | 0.450 |

Data shown are number (%) or median (interquartile range).

These patients overall were tested for humoral immunity function. However, there was no significant difference in IgG, IgM, IgA, C3, C4 and the percentage of IgE > 100 IU/ml between the death group and the survival group (Table 3).

**Table 3.** Humoral immunity in survival and death groups

|  | Normal range | All patients | Survival | Death | *P* |
| --- | --- | --- | --- | --- | --- |
| Subjects, n |  | 85 | 71 | 14 | — |
| IgG, g/L | 8-16 | 11.80(10.20,13.45) | 10.00(11.70,13.40) | 12.20(10.60,14.30) | 0.362 |
| IgM, g/L | 0.4-3.45 | 0.88(0.68,1.07) | 0.86(0.67, 1.09) | 0.93(0.72,1.16) | 0.358 |
| IgA, g/L | 0.76-3.90 | 2.62(1.80,3.16) | 2.55(1.81,3.01) | 3.30(1.68,4.50) | 0.211 |
| IgE≥100 IU/mL, n(%) | <100 | 27(31.76%) | 23(32.39%) | 4(28.57%) | 0.779 |
| C3, g/L | 0.81-1.60 | 1.030(0.913,1.125) | 1.040(0.913,1.150) | 0.987(0.819,1.048) | 0.158 |
| C4, g/L | 0.1-0.4 | 0.243(0.186,0.317) | 0.243(0.187,0.304) | 0.217(0.169,0.355) | 0.873 |

Univariate and multivariate logistic regression analyses were performed to determine which admission factors were independently associated with mortality (Table 4). After controlling for other variables, it was found that only the admission neutrophil count was an independent risk factor for mortality (adjusted OR 1.672; 95% confidence interval [CI] 1.172-2.385; *P* = 0.005).

**Table 4.** Univariate and multivariate logistic regression to identify risk factors at admission related to mortality

|  | Univariate |  | Multivariate |  |
| --- | --- | --- | --- | --- |
|  | Unadjusted OR (95% CI) | *P* | Adjusted OR (95% CI) | *P* |
| Neutrophil count | 1.775(1.351- 2.333) | 0.000 | 1.672 (1.172- 2.385) | 0.005 |
| Lymphocyte count | 0.185 (0.040- 0.867) | 0.032 | 0.022 (0.000- 4.503) | 0.160 |
| CD3+ count | 0.997 (0.994- 0.999) | 0.015 | 1.006 (0.997- 1.015) | 0.212 |
| CD4+ count | 0.993 (0.988- 0.998) | 0.004 | 0.996 (0.979- 1.013) | 0.629 |
| CRP > 60 mg/L | 8.730 (2.028- 34.514) | 0.002 | 4.556 (0.572- 36.297) | 0.152 |
| PaO<sub>2</sub>/FiO<sub>2</sub> | 0.983 (0.971- 0.994) | 0.004 | 0.999(0.988-1.010) | 0.797 |

CI- confidence interval; CRP- C-reactive protein; OR- odds ratio.

### ROC curve analyses of mortality

Areas under the ROC curves (AUCs) as predictors of mortality, determined from the analyses of blood parameters at baseline, were calculated and compared (Table 5). The AUCs showed that the following could predict death due to COVID-19: leukocyte count; neutrophil percentage; neutrophil count; lymphocyte percentage; lymphocyte count; CD3+ %; CD3+ count; CD4+ %, CD4+ count and PaO_2_/FiO_2_.

**Table 5.** AUCs determined from ROC curves for predictors of mortality due to COVID-19.

|  | AUC (95% CI) | *P* |
| --- | --- | --- |
| Leukocyte count | 0.874 (0.774- 0.975) | 0.000 |
| Neutrophil % | 0.918 (0.859-0.976) | 0.000 |
| Neutrophil count | 0.909 (0.835- 0.984) | 0.000 |
| Lymphocyte % | 0.916 (0.856-0.977) | 0.000 |
| Lymphocyte count | 0.768 (0.620- 0.916) | 0.002 |
| CD3+ Count | 0.729 (0.593-0.865) | 0.007 |
| CD4+ count | 0.786 (0.687-0.886) | 0.001 |
| PaO<sub>2</sub>/FiO<sub>2</sub> | 0.816 (0.665-0.966) | 0.000 |

### Dynamic profile of blood parameters after 2 weeks hospitalization in survivors

In the survival group (n = 71), after treatments of symptoms, cough, shortness of breath, and dyspnea were relieved to varying degrees, and the chest CT gradually improved. Four patients were well enough for discharge within 2 weeks after admission. The remaining hospitalized survivors (n=67) underwent blood tests, 2 weeks after treatment. Lymphocyte count and neutrophil count return to normal range in most patients. However, in 13 of the 67 surviving patients after 2 weeks of treatment, the lymphocyte count was still below the lower limit of normal (1.1 × 10^9^/L). In addition, in 4 of the 67 surviving patients after 2 weeks of treatment, the neutrophil count was above the higher limit of normal (6.3 × 10^9^/L).

In 53 patients who tested cellular immunity of the survival group after 2 weeks of hospitalization, the count of lymphocyte subsets returned to normal range in most patients (Table 7). However, in 13 of these 53 patients, the CD3+ count remained lower than the lower limit of the normal reference value (723/µL); in 10 of these 55 patients, the CD4+ count was still lower than the lower limit of the normal reference value (404/µL).

**Table 6.** Blood parameters in survival group at admission and 2 weeks after admission *

|  | Normal range | After 2 weeks (n=67) |
| --- | --- | --- |
| Leukocyte count, $\times 10^9/L$ | 3.5-9.5 | 5.32 (4.47,6.26) |
| >9.5, n (%) |  | 3 (4.48%) |
| 3.5-9.5, n (%) |  | 61 (91.04%) |
| <3.5, n (%) |  | 3 (4.48%) |
| Neutrophil percentage, % | 40-75 | 59.50 (56.60, 63.80) |
| Neutrophil count, $\times 10^9/L$ | 1.8-6.3 | 3.10 (2.56, 3.92) |
| >6.3, n (%) |  | 4 (5.97%) |
| 1.8-6.3, n (%) |  | 60 (89.55%) |
| <1.8, n (%) |  | 3 (4.48%) |
| Lymphocyte percentage, % | 20-50 | 27.20 (23.30, 31.10) |
| Lymphocyte count, $\times 10^9/L$ | 1.1-3.2 | 1.46(1.25,1.70) |
| >3.2, n (%) |  | 0 |
| 1.1-3.2, n (%) |  | 54 (80.60%) |
| <1.1, n (%) |  | 13 (19.40%) |
| Hemoglobin, g/L | 115-150 | 112.00 (104.00,125.00) |
| Platelets, $\times 10^9/L$ | 125-350 | 243.00 (194.00, 303.00) |

\* n = 67. Data shown are number (%) or median (interquartile range).

**Table 7.** Cellular immunity in survival group at admission and 2 weeks after admission *

|  | Normal range | After 2 weeks (n = 53) |
| --- | --- | --- |
| CD3+ percentage, % | 56-86 | 68.50(61.44, 75.51) |
| CD3+ count, /$\mu L$ | 723-2737 | 863.00(717.00, 1174.50) |
| <723/$\mu L$, n (%) | | 13 (24.53%) |
| CD4+ percentage, % | 33-58 | 41.61 (34.87, 46.65) |
| CD4+ count, /$\mu L$ | 404-1612 | 544.00 (448.00, 692.50) |
| <404/$\mu L$, n (%) | | 10 (18.87%) |
| CD8+ percentage, % | 13-39 | 24.08(18.30, 29.74) |
| CD8+ count, /$\mu L$ | 220-1129 | 301.00(240.50, 418.50) |
| <220/$\mu L$, n (%) | | 10(18.87%) |
| CD4+/CD8+ ratio | 0.9-2.0 | 1.73(1.35, 2.34) |
| <0.9, n (%) |  | 2(3.77%) |
| CD19+ percentage, % | 5-22 | 11.03(8.41,13.35) |
| CD19+ count, /μL | 80-616 | 140.00(103.00, 202.50) |
| <80/μL, n (%) | | 5(9.43%) |
| CD16+56+ percentage, % | 5-26 | 18.22(10.94, 22.09) |
| CD16+56+ count, /μL | 84-724 | 217.00(143.50, 328.50) |
| <84/μL, n (%) | | 2(3.77%) |

\* n = 53. Data shown are number (%) or median (interquartile range).

## DISCUSSION

Since December 2019, COVID-19 has been prevalent in Wuhan, China. It is highly contagious through human-to-human transmission, and in severe cases can lead to death [15]. Patients with severe disease may have respiratory manifestations such as fever, cough, shortness of breath, dyspnea, and hypoxemia. Chest CT examination of the patients shows predominant ground glass opacities mixed with consolidations, mainly peripheral or combined peripheral and central distributions, bilateral and lower lung zones being mostly involved [16]. In addition, COVID-19 can attack and lead to the collapse of the immune system in the critically ill. Serious inhibition of the immune system may promote replication of the virus and its spread throughout the body, damaging the function of multiple organs.

The present retrospective study of patients with COVID-19 found no statistical association between age and mortality, and none between the interval from onset to hospitalization and mortality. The percentage of men and rate of concomitant disease were higher in the patients who died compared with survivors, but the difference was not significant, perhaps due to the smallness of the sample.

However, the admission (baseline) leukocyte count, neutrophil percentage, and neutrophil count of the death group were all significantly higher than that of the survivors. Chang et al. [2] reported that an initial neutrophil count >7000/mL was an independent risk factor for death in patients with SARS. Singapore scholars also concluded that neutropenia predicted poor prognosis in SARS [3]. The leukocyte counts of severe patients with COVID-19 were significantly higher than those of non-severe patients [8-10]. For the present patients with COVID-19, we found that neutrophil count was above the higher limitation of normal range in 12(85.71%) dead patients and an initial elevated neutrophil count was an independent risk factor of mortality. This significant elevation of neutrophil count in the death group, may reflect a strong inflammatory response toward viral infection, or a possible combination with bacterial infection. The specific mechanism needs further study.

The report of Wang et al. [9] on patients with COVID-19 showed that the lymphocyte count continued to decrease with progression, unto death. The latest researches showed that the severe COVID-19 cases had significantly lower lymphocyte counts compared with non-severe COVID-19 cases [8-13]. The present study also determined that more than half of the patients (53/85, 62.35%) had lower lymphocyte count than the lower limitation of normal, especially in those who died (12/14, 85.7%). And we found that the lymphocyte counts in dead group were significantly lower than those in survival group. It is not clear whether there are other mechanisms for lymphocyte decline in COVID-19, other than the destruction of lymphoid organs by viral attack. Lymphopenia in patients with SARS before hormone therapy may be induced by a stress mechanism involving the hypothalamus-pituitary-adrenal axis, due to elevation of the cortisol level [17].

Lymphocyte subsets have an important role in humoral and cellular immunity against viral infection. The prognosis of various viral diseases is closely linked to cellular immune function, especially T lymphocyte function. Qin et al. [9] found that number of helper T cells, suppressor T cells and regulatory T cells decreased in patients with COVID-19. Wang et al. [13] reported that CD4+ T cells, CD8+ T cells, B cells, and natural killer (NK) cells decreased in patients with COVID-19, and were significantly lower in severe cases than mild cases. Chen et al. [8] showed that CD4+T and CD8+T cells reduced in nearly all patients with COVID-19, and severe cases had a lower level than moderate cases. In the present study of patients with COVID-19, the baseline median values of CD3+, CD4+, and CD8+ counts were lower than the lower limit of the normal range. What is more noteworthy is that the CD3+ and CD4+ count levels in the patients who died was significantly lower than that of the survivors. The baseline CD8+, CD19+, and CD16+56+ counts in those who died were also lower compared with the survivors, although the differences between the two groups were not statistically significant.

A study reported the suppression of cellular immunity in patients who died of viral pneumonia [18]. One study found that the counts of peripheral CD4+ and CD8+ T cells were substantially lower than normal in a 50-year-old man who died from COVID-19 infection [14]. We speculate that the decrease in number of lymphocyte subsets in COVID-19 may be caused by an attack of the immune system by SARS-CoV-2.

Lam et al. [19] reported that CD3+, CD4+, CD8+, and natural killer cell counts were promising predictors for intensive care unit admission in patients with SARS. In the present study, the ROC analysis determined that admission leukocyte count, neutrophil percentage, neutrophil count, lymphocyte percentage, and lymphocyte count were able to predict the mortality of COVID-19 patients. In terms of cellular immune function, CD3+%, CD3+ count, CD4+%, CD4+ count and PaO_2_/FiO_2_ could also predict mortality. Clinicians should pay attention to these parameters when caring for these patients.

At present, there is no study about humoral immunity function of patients with COVID-19. We found that the difference of IgG, IgM, IgA, C3, C4 and the percentage of IgE > 100 IU/ml was not significant between the survivors and non-survivors. The baseline platelet level in patients who died was lower than that of the survivors. Reference to other virus researches, this may be due to platelet destruction caused by the virus, or inhibition of the production of platelets in bone marrow via viral infection [20], or platelet consumption caused by the formation of thrombus at the injured site of lung [21,22]. The hemoglobin level in the dead patients is higher than that in the surviving patients, which may be due to the severe lung injury in the dead patients and the irritant increase of hemoglobin caused by hypoxemia.

Compared with admission, after 2 weeks of hospitalization the blood routine examination of 67 patients in the group who survived showed neutrophil and lymphocyte count of most patients returned to the normal range. It is worth noting that the lymphocyte count levels in 13 of the 67 surviving patients did not rise to the lower limitation of normal levels (1.1 × 10^9^/L) after 2 weeks of treatment, suggesting that the immune function of these patients had not returned to normal. In addition, the lymphocyte subsets of some patients also did not return to the normal range. The positive rate of SARS-CoV-2 RNA in nasopharyngeal swabs was not satisfactory. When considering the hospital discharge of patients with COVID-19, improved symptoms, obvious absorption in the chest CT, and consecutive negative nucleic acid tests are important. However, we recommend in addition that lymphocyte count and cellular immune function tests should be conducted to ensure that the disease has been effectively controlled. A previous study of patients with SARS showed that, after two years, the counts of peripheral blood lymphocytes and subsets, comprising lymphocytes, CD4+, CD8+, and NK cells (CD16+56+), had remained lower than that of the healthy controls [23]. In patients with COVID-19, changes in lymphocytes and the lymphocyte subsets during recovery need to be studied.

There are several limitations to the present work. Firstly, it is a single-center retrospective with a small sample size. However, to the best of our knowledge, it is the first to analyse the dead and survival patients with COVID-19 the routine blood and immune function indexes. Secondly, in surviving patients the selected timepoints for examining blood indexes were at admission and 2 weeks after beginning hospitalized treatment, which may not accurately reflect the continuous dynamic changes in leukocytes, neutrophils, lymphocytes, and lymphocyte subsets. Finally, the number of patients given immune function tests who later died is relatively small; and the only evaluations of immune function were for cellular immunity and humoral immunity.

Of note, the mortality rate in this study was relatively high. One study of COVID-19 patients reported a death rate of 6 patients (4.3%) among 138 patients [6]. In another study, the mortality rate of COVID-19 was 11.0% [24]; while a recent large-sample and multicenter study showed that only 1.36% of patients succumbed [25]. We should point out that all the patients in the present study who died did so before 11 February 2020. The higher mortality rate was likely the result of the heightened severity of disease of admitted patients at this early stage of the epidemic in China. In addition, the treatment area was a temporary structure, expediently built, without a well-equipped Intensive Care Unit, and the medical conditions were insufficient for the need.

## Conclusions

In this population of COVID-19 patients, the admission leukocyte count, neutrophil percentage and neutrophil count in those who died within 28 days were all higher than that of the survivors, and the lymphocyte count, CD3+ count and CD4+ count was lower. The neutrophil count was independently associated with mortality. Leukocyte, neutrophil, lymphocyte, CD3+ and CD4+ count at admission could predict the mortality due to COVID-19.

## Data Availability

The data used to support the findings of this study are available from the corresponding author upon request.

## Contributors

FX conceived the study, coordination and finalized the manuscript. YF participated in the design, data acquisition, database management, statistical analysis and coordination. YS participated in statistical analysis and the manuscript draft. YY and YW participated in data acquisition. SL participated in the database management. All authors read and approved the final manuscript. FX takes responsibility for the article as a whole.

